# TERT promoter mutation analysis for blood-based diagnosis and monitoring of gliomas

**DOI:** 10.1101/2020.09.03.20135236

**Authors:** Koushik Muralidharan, Anudeep Yekula, Julia L. Small, Zachary S. Rosh, Keiko M. Kang, Lan Wang, Spencer Lau, Hui Zhang, Hakho Lee, Chetan Bettegowda, Michael R. Chicoine, Steven N. Kalkanis, Ganesh M. Shankar, Brian V. Nahed, William T. Curry, Pamela S. Jones, Daniel P. Cahill, Leonora Balaj, Bob S. Carter

## Abstract

Liquid biopsy offers a minimally invasive tool to diagnose and monitor the heterogeneous molecular landscape of tumors over time and therapy. Gliomas have been challenging to sample with blood-based liquid biopsy due to low levels of circulating tumor-derived nucleic acid, and prior efforts have shown specific point mutations in less than 5% of glioma patients. Detection of TERT promoter mutations (C228T, C250T) in cfDNA has been successful for some systemic cancers but has yet to be demonstrated in gliomas, despite the high prevalence of these mutations in glioma tissue (>60% of all tumors). Here, we developed a novel digital droplet PCR (ddPCR) assay, that incorporates features to improve sensitivity and allows for the simultaneous detection and longitudinal monitoring of two TERT promoter mutations (C228T and C250T) in cfDNA from the plasma of glioma patients. In baseline performance in tumor tissue, (*n* = 157 samples), the assay had perfect concordance with an independently performed clinical pathology laboratory assessment of TERT promoter mutations in the same tumor samples (95% CI 94%-100%). Extending to matched plasma samples, we detected TERT mutations in both discovery and blinded multi-institution validation cohorts with an overall sensitivity of 62.5% (95% CI 52%-73%) and a specificity of 90% (95% CI 80%-96%) compared to the gold standard tumor tissue-based detection of TERT mutations. Upon longitudinal monitoring in 5 patients, we report that peripheral TERT mutant allele frequency reflects the clinical course of the disease with levels decreasing after surgical intervention and therapy and increasing with tumor progression. Our results demonstrate the feasibility of detecting circulating cfDNA TERT promoter mutations in glioma patients with clinically relevant sensitivity and specificity using a novel, enhanced ddPCR approach. The functionality of this assay also suggests a new clinical role for plasma-based cfDNA analysis to complement tissue-based histopathologic and molecular characterization for diagnosis and spatiotemporal monitoring of brain tumors.

## Introduction

Liquid biopsy for the detection and monitoring of brain tumors is of significant clinical interest. Upon clinical presentation, patients typically undergo imaging followed by biopsy alone or biopsy with resection for diagnosis and determination of histopathological classification^1^. While tissue biopsy is invasive and can, in some cases, be high risk, liquid biopsy can offer a less invasive sampling approach that still affords significant clinical information for diagnosis. In addition, liquid biopsy can be performed more frequently to allow for longitudinal treatment monitoring. The detection of glioma mutations via liquid biopsy of cerebrospinal fluid has been demonstrated^2-6^; however, the ability to detect mutations in cell free DNA (cfDNA) in plasma with similar sensitivities has been limited to date^5,7,8^.

TERT promoter mutations are amongst the most common genetic alterations in glioma^6,9,10^.The two most common mutations are C228T (45% of all TERT promoter mutations) and the C250T (15% of all TERT promoter mutations), occurring 124 and 146 bp respectively upstream of the TERT transcriptional start site^11^. These mutations are typically heterozygous, mutually exclusive, and due to the GC-rich nature of the promoter, yield an identical 11bp ‘CCCGGAAGGGG’ ETS/TCF binding motif that is associated with increased transcriptional activity of TERT, and therefore increased telomerase activity^12,13^. Somatic mutations in TERT promoter are common across all grades of gliomas with a frequency of 8% in pilocytic astrocytomas (grade I), 15% in diffuse astrocytomas (grade II), 45% in oligodendrogliomas (grade II), 54% in anaplastic oligodendrogliomas (grade III) and 62% in glioblastoma multiforme (grade IV)^9,10,14,15^. Additionally, presence of TERT mutation is a poor prognostic marker in these glioma subtypes^9,10,15^. To date, PCR amplification of TERT has proven technically difficult due to the high GC content in the promoter region (>80%)^16,17^. To address these issues, several additive and non-additive based approaches including Q-sol, 7-deaza-dGTP and locked nucleic acid (LNA)-enhanced probes have been previously reported^11,18-20^.

In this study, we have developed a novel ddPCR probe-based assay that can detect two TERT promoter mutations (C228T and C250T) in the plasma of glioma patients with dramatically improved specificity and sensitivity (**Fig. 1a**). As a baseline, we showed that performance of the assay in tumor tissue compared perfectly to independent determined clinical pathology laboratory results, with ability to discriminate between C250T and C228T mutant alleles. In addition, using an unbiased, R-based statistical analysis of ddPCR data, we show the detection of TERT promoter mutations in an analysis of tumor tissue matched plasma samples and healthy control samples in both discovery (n=83) and blinded multi-institutional validation (n=74) cohorts with an overall (n=157) sensitivity of 62.5% and specificity of 90%. Finally, we show the potential of our assay for use in longitudinal monitoring, showing rising and falling peripheral TERT levels in concordance with responses to glioma treatment and disease recurrence.

**Figure 1.**
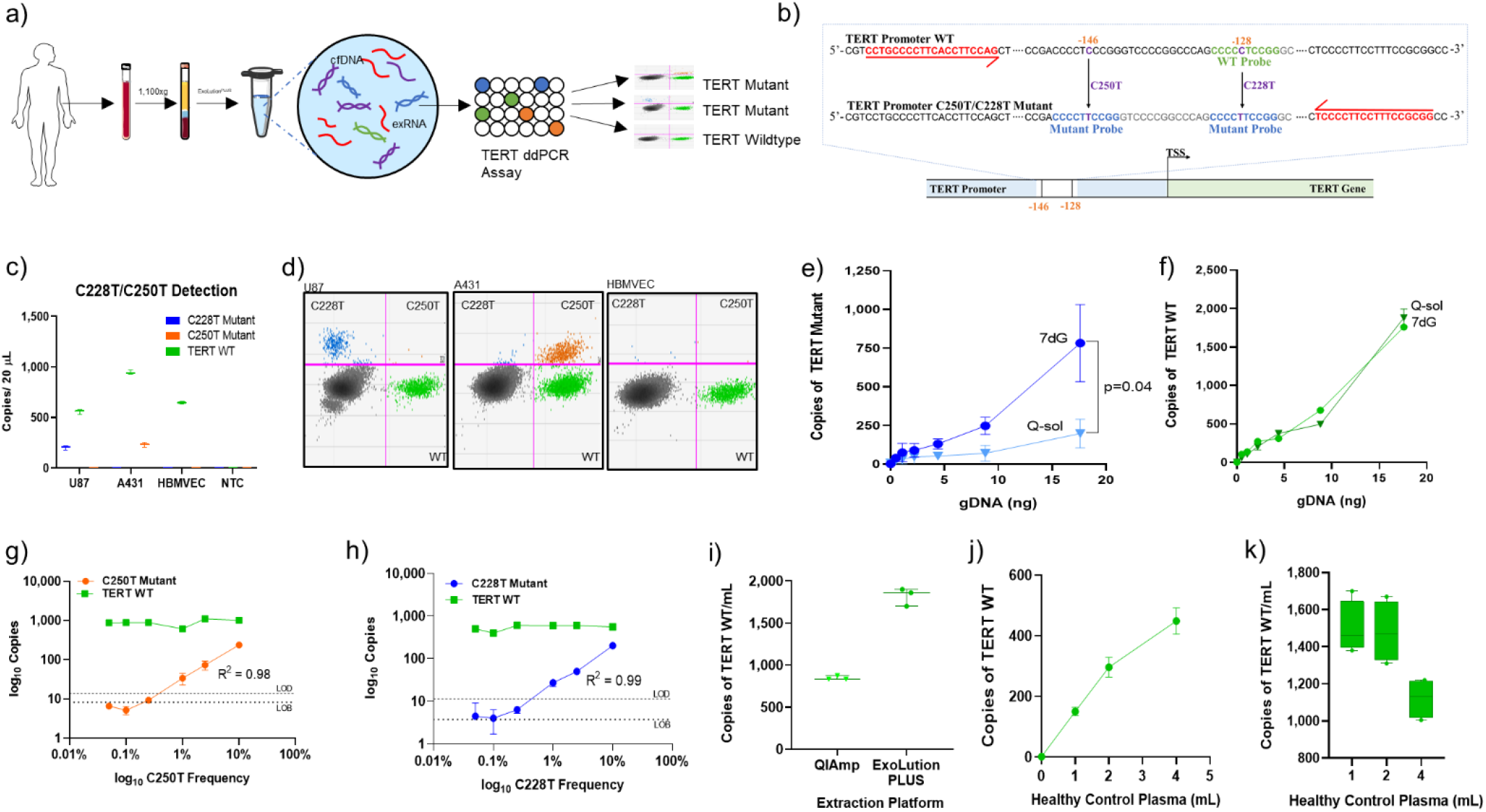
Overview and Optimization of TERT Promoter Mutation ddPCR Assay. **(a)** Schematic depicting experimental workflow, including isolation of plasma, extraction platform, and TERT Promoter ddPCR readout. **(b)** TERT promoter nucleotide sequence illustrating forward and reverse primers as well as probes specific to each mutation. **(c)** Absolute quantification of TERT mutant copies from equal inputs of genomic DNA from U87 (C228T), A431 (C250T), HBMVEC (WT) cell lines. **(d)** 2D amplitude plots indicate the mutant and wildtype populations for each specific mutation and cell line. Y-axis indicates positivity in the mutant channel, and the X-axis indicates positivity in the wildtype channel. Events positive for both channels are shown in the upper right corner of the 2D amplitude plot. Both mutant and wildtype probes can bind the C250T mutation, due to the location of the point mutation (positive in the lower and upper right corner). However, either the wildtype probe or the mutant probe can exclusively bind to the C228T mutation (positive in the upper Left and lower right corner). Background signal is seen in the lower left corner. Serial dilutions of genomic DNA (gDNA) from A431 cells are used as templates for TERT ddPCR assay using 7-deaza-dGTP Q-sol as an additive. Copies per 20μL of TERT Mutant **(e)** and TERT WT **(f)** are plotted against input (in nanograms) of A431 gDNA. Serial dilutions of gDNA from TERT Mutant cell lines **(g)** U87 (C228T) and **(h)** A431 (C250T) were diluted in a constant background of TERT WT HBMVEC gDNA from 10% mutant allele frequency to 0% mutant allele frequency. Copies per 20 μL of TERT Mutant and TERT WT are plotted against mutant allele frequency. Limit of detection (LOD, dashed line) is plotted, defined as 2 standard deviations over the mean frequency abundance obtained at 0% when only TERT WT DNA was used as input. Limit of blank (LOB, dotted line) is plotted, defined as the highest apparent mean frequency abundance expected to be found when replicates of a blank sample containing no TERT Mutant copies are tested. **(i)** cfDNA was extracted from 2 mL of healthy control plasma using the QIAmp circulating nucleic acid kit (QIAGEN) and the ExoLution PLUS extraction kit (Exosome Diagnostics). 2 μL of cfDNA was used as input for absolute quantification of TERT WT cfDNA. Copies/mL were calculated using the formula described in Methods. (**j,k**) cfDNA was extracted from 1 mL, 2mL, and 4 mL of healthy control plasma using the ExoLution PLUS kit (Exosome Diagnostics). 2 μL of cfDNA was used as input for absolute quantification of TERT WT cfDNA. Copies per 20 μL **(j)** and Copies/mL **(k)** are plotted against the amount of healthy control plasma used for reaction.

## Results

### Assay Design and Optimization

The most common TERT promoter mutations, C228T and C250T, are heterozygous and mutually exclusive, but both mutations result in the generation of an 11-bp identical sequence, 5’-CCCCTTCCGGG-3’. We used a 10-bp LNA mutant probe to simultaneously detect both mutations and an LNA wildtype probe complementary to the C228 locus (**Fig. 1b**). Assay specificity for each mutation was established using U87 (C228T mutant), A431 (C250T mutant), and HBMVEC (TERT WT) gDNA (**Fig. 1c**). For inputs with mutant allele frequencies (MAF) greater than 10%, 2D amplitude analysis can distinguish between C250T/C228T mutations (**Fig. 1d**). TERT promoter mutations are situated in a GC-rich region resistant to amplification that hinders assay performance. To stabilize amplification, we compared Q-sol additive to 7-deaza-dGTP (7dG), a modified nucleotide that inhibits secondary structure formation^17^. TERT assay performance with 7dG was superior to Q-Sol (1.8-3.9-fold increase, p=0.04) with higher absolute mutant detection and comparable WT detection (**Fig. 1e-f**). We report analytical parameters including limit-of-detections (LODs) of 0.27% MAF (C250T) and 0.42% (C228T) MAF and limit of blanks of 0.02% (C250T) and 0.04% MAF (C228T; **Fig. 1g-h; Extended Data Fig. 1**). In a comparison of extraction platforms to optimize TERT recovery, a two-fold increase in TERT WT (p=0.001) was seen while using ExoLution PLUS in comparison to the QiAmp Circulating Nucleic Acid kit (Qiagen) (**Fig. 1i**). Two mL of plasma was determined to be the optimal input for recovery as a function of copies of TERT WT per mL of plasma (**Fig. 1j-k**).

**Figure 2.**
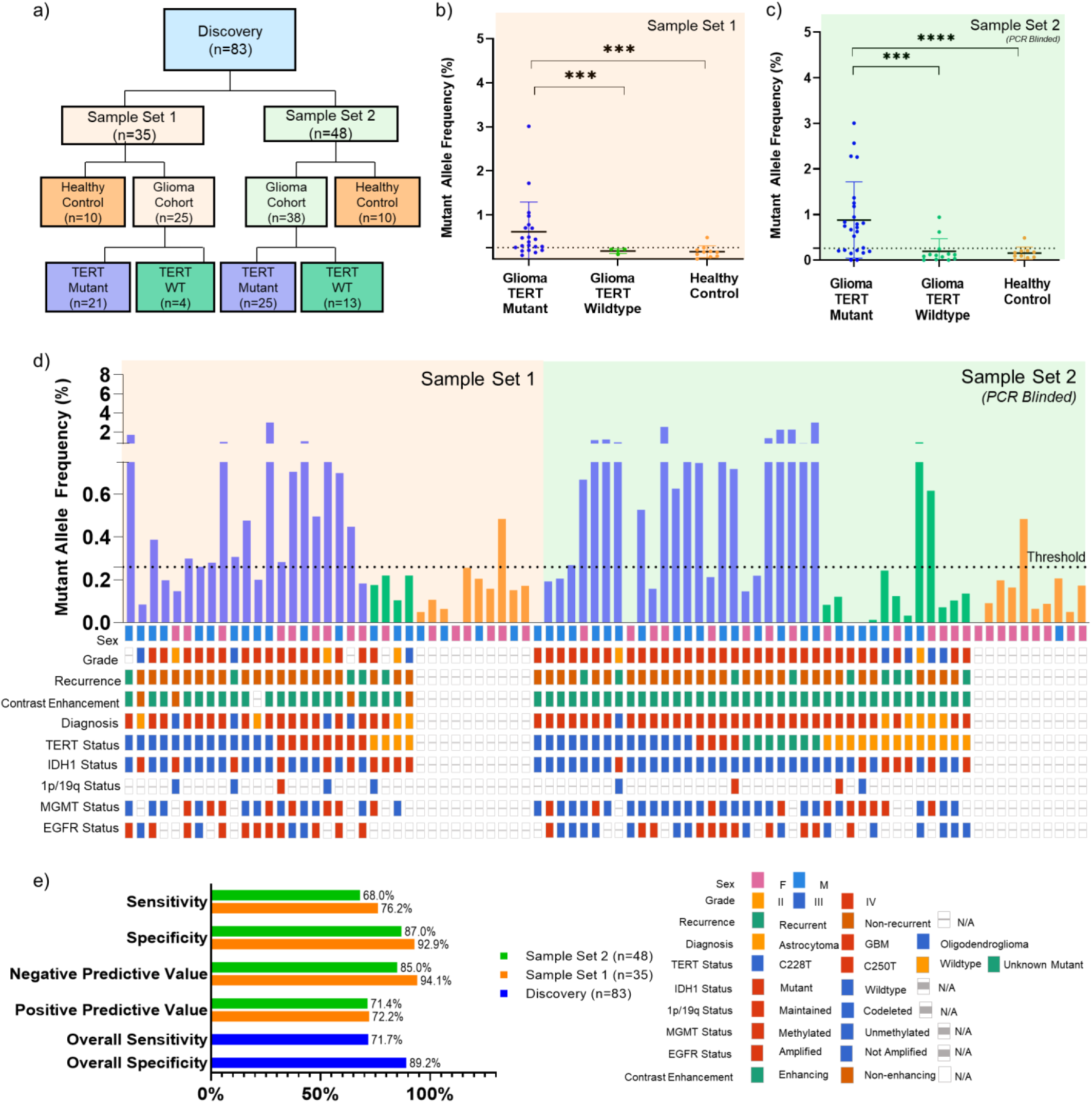
Detection of TERT Promoter Mutation in Plasma of Discovery Cohort. **(a)** CONSORT diagram depicting patient cohorts and overall study design. **(b)** cfDNA from 2 mL of matched patient plasma from patient cohort 1 (21 TERT Mutant and 4 TERT WT; n=25) and healthy control (TERT WT; n=10) was used as input for absolute quantification of TERT mutant (C228T or C250T) and wildtype copies. **(c)** Plasma samples from PCR blinded sample set 2 (25 TERT Mutant and 13 TERT WT; n=38) and healthy control (TERT WT; n=10) was used as input for absolute quantification of TERT mutant (C228T or C250T) and analyzed using parameters established in sample set 1. All data is shown in MAF, calculated using the formula described in **Methods and Extended Data Fig. 2**. MAF of TERT Mutant for plasma samples are plotted against cohort sub-classification. Dotted line indicates a threshold of 0.26% MAF, used to designate samples as TERT Mutant positive or negative. **(d)** MAF of TERT Mutant are graphed according to sample number. Oncoprint depicting the genomic landscape of each sample are plotted underneath. **(e)** Contingency tables were constructed from data obtained, and sensitivity, specificity, negative predictive value and positive predictive value were calculated and graphed above. Overall sensitivity and specificity across both sample sets(n=83) are also reported.

### Development of R-based Analysis for TERT Promoter ddPCR Analysis and Cohort Design

To standardize gating without operator bias, we sought to mathematically define and automate the gating strategy. Our gating strategy is based on the algorithm used by the R-program *ddPCR*, which defines empty droplets as those that lie within 7 standard deviations above the mean amplitude of channel 1^21^. To determine the optimal gating strategy, we gated continuously from 4-8 standard deviations above the mean amplitude of channel 1 and 2-6 standard deviations above the mean amplitude of channel 2 in increments of 0.05, with pseudocode provided in **Extended Data Fig. 2**. Bootstrapping with 1000 replicates was used to determine a threshold, and a list of gating strategies with threshold values was generated to ensure that specificity was greater than or equal to 90%. We present one such gating strategy that maximizes the sum of discovery sensitivity, validation sensitivity, and overall sensitivity. This gating strategy gates at 7.15 standard deviations above the mean of channel 1 amplitude, 2.95 standard deviations above the mean of channel 2 amplitude, with a threshold of 0.26% MAF **(Extended Data Fig. 2**). This is in accordance with prior literature, which suggests that positive droplets lie 7 standard deviations above the mean amplitude of channel 1^21^. Notably, the threshold calculated by the program, 0.26 % MAF, was equivalent to the experimentally determined LOD (MAF) of the TERT Promoter ddPCR Assay using cell line derived gDNA (**Fig. 1g**).

### Detection of TERT Promoter Mutations in cfDNA from Discovery and Blinded Validation Cohort

We selected a cohort (*n* = 157) of molecularly characterized glioma patients (*n* = 114), non-tumor patients with enhancing lesions on MRI (*n* = 9), and age matched healthy controls (*n* = 33). Our patient population spanned tumor diagnosis (20% astrocytoma, 7% oligodendroglioma, 72% glioblastoma, 1% gliosarcoma); grade (6% II; 15% III, 71% IV, 8% not reported) and molecular characteristics: IDH1 mutant (19%), TERT mutant (45% C228T; 15% C250T), 1p19q codeletion (7%), EGFR amplified (20%), MGMT methylated (33%) (**Table 1; Fig. 2**). The study population was randomly assigned to either a discovery cohort (*n* = 83) or a blinded multi-institutional validation cohort (*n* = 74). To assess the clinical performance of the assay, tumor tissue and matched plasma samples were analyzed for the presence of the TERT mutations (**Extended Data Fig. 3**).

**Table 1.** Patient Demographics. IQR (Interquartile Range). N/A = Not applicable.

|  | Discovery Set 1 | % | Discovery Set 2 | % | Blinded Multi-Institution Validation Cohort | % |
| --- | --- | --- | --- | --- | --- | --- |
| **Total** | 35 |  | 48 |  | 74 |  |
| Disease Group Age median, IQR | 50 (43-64) |  | 59.5 (50-68.5) |  | 56 (46-66.5) |  |
| Healthy Group Age median, IQR | 31.5 (26-52.75) |  | 26.5 (24-29.75) |  | 55.5 (48.5-63.3) |  |
| Non-Tumor Group Age median, IQR | N/A |  | N/A |  | 66 (62-67) |  |
| **Sex** |  |  |  |  |  |  |
| Male | 16 | 45.7 | 25 | 52.1 | 46 | 62.2 |
| Female | 19 | 54.3 | 23 | 47.9 | 28 | 37.8 |
| **Race, n (%)** |  |  |  |  |  |  |
| White | 31 | 88.6 | 43 | 89.6 | 65 | 87.8 |
| Asian | 1 | 2.9 | 2 | 4.2 | 3 | 4.1 |
| Black | 0 | 0.0 | 0 | 0.0 | 3 | 4.1 |
| Other | 2 | 5.7 | 1 | 2.1 | 0 | 0.0 |
| Unavailable | 1 | 2.9 | 2 | 4.2 | 3 | 4.1 |
| **Ethnicity, n (%)** |  |  |  |  |  |  |
| Hispanic | 1 | 2.9 | 2 | 4.2 | 0 | 0.0 |
| Non-Hispanic | 30 | 85.7 | 40 | 83.3 | 55 | 74.3 |
| Unavailable | 4 | 11.4 | 6 | 12.5 | 19 | 25.7 |
| **WHO Grade, n (%)** |  |  |  |  |  |  |
| II | 3 | 8.6 | 2 | 4.2 | 2 | 2.7 |
| III | 3 | 8.6 | 4 | 8.3 | 10 | 13.5 |
| IV | 16 | 45.7 | 32 | 66.7 | 33 | 44.6 |
| N/A | 13 | 37.1 | 10 | 20.8 | 29 | 39.2 |
| **Disease Status** |  |  |  |  |  |  |
| Newly Diagnosed | 21 | 60.0 | 30 | 62.5 | 39 | 52.7 |
| Recurrent | 4 | 11.4 | 8 | 16.7 | 18 | 24.3 |
| N/A |  |  |  |  | 17 | 23.0 |
| **Medications, n (%)** |  |  |  |  |  |  |
| anticonvulsants (levetiracetam, clonazepam, lacosomide, carbamazepine, lamotrigine) | 16 | 45.7 | 13 | 27.1 | 30 | 40.5 |
| steroids (dexamethasone) | 7 | 20.0 | 10 | 20.8 | 18 | 24.3 |
| blood thinner (aspirin, coumadin) | 3 | 8.6 | 4 | 8.3 | 9 | 12.2 |
| prior radiation therapy | 1 | 2.9 | 5 | 10.4 | 2 | 2.7 |
| prior chemotherapy | 2 | 5.7 | 8 | 16.7 | 4 | 5.4 |
| clinical trial participant | 1 | 2.9 | 1 | 2.1 | 6 | 8.1 |

In tissue, we demonstrated that our assay and the CLIA certified Solid SNAPSHOT assay^22^ used in the MGH Department of Pathology, detected TERT positive tumors with 100% concordance across 97 tested samples for the presence of the TERT promoter mutation. (**Fig 2a, Extended Data Fig. 3**). In one patient, using parallel tumor tissue aliquots, our assay detected the C250T mutation while the SNAPSHOT assay detected the C228T mutation.

Plasma samples (Total n=83; TERT mutant n=46; TERT WT n=17; healthy controls n=20) were analyzed and gated (**Extended Data Fig. 2**) in two separate discovery sample sets (**Fig. 2a**). In sample set 1, we report positivity in 16 of 21 (76%) TERT mutant samples, and in 1 of 14 (7%) WT control samples (**Fig. 2b**), with a sensitivity of 76.19% (CI, 52.83 - 91.78), specificity of 92.86% (CI, 66.3 - 99.82), PPV of 94.12% (CI, 70.45 - 99.08), and NPV of 72.22% (95% CI, 54.41 - 85.00; **Fig. 2e**). In sample set 2 (PCR blinded), we report positivity in 17 of 25 (68%) of TERT mutant samples, and in 3 of 20 (15%) of WT control samples (**Fig 2c**), with a sensitivity of 68.00% (CI, 46.50 - 85.05), specificity of 86.96% (CI, 66.41 - 97.22), PPV of 85.00% (CI, 65.6 - 94.39), NPV of 71.43% (CI, 58.01 - 81.89; **Fig 2d**). Overall, in the discovery cohort, we report positivity in 33 of 46 (72%) TERT mutant samples, and in 4 of 37 (11%) TERT WT samples (**Fig 2d**), with a sensitivity of 71.74% (CI, 56.54 - 84.01) and specificity of 89.19% (CI, 74.58 - 96.97).

To validate assay performance in multi-institutional samples, we analyzed a blinded cohort (Total n=74; TERT mutant n= 41 (Henry Ford Hospital n= 12; Washington University n=2; Massachusetts General Hospital n=27); TERT WT n=17; healthy control n=14, CNS disease non-glioma n=9) using the parameters previously described (**Fig. 3a, Extended Data Fig. 2**). The CNS disease non-tumor samples are from patients who either had a non-malignant contrast enhancing mass on MRI, such as a demyelinating lesion or fungal abscess, that was initially suspected as glioma or other non-tumor conditions such as normal pressure hydrocephalus. In matched plasma, we report positivity in 22 of 42 (52%) confirmed mutant samples and 3 of 33 (9%) confirmed wild-type with a sensitivity of 52.38% (CI, 36.42%-68.00%), specificity of 90.91% (CI, 75.67% to 98.08%), PPV of 88% (CI, 70.59% to 95.73%), NPV of 72% (CI, 51.76% to 67.71%) (**Fig. 3b-d**).

**Figure 3.**
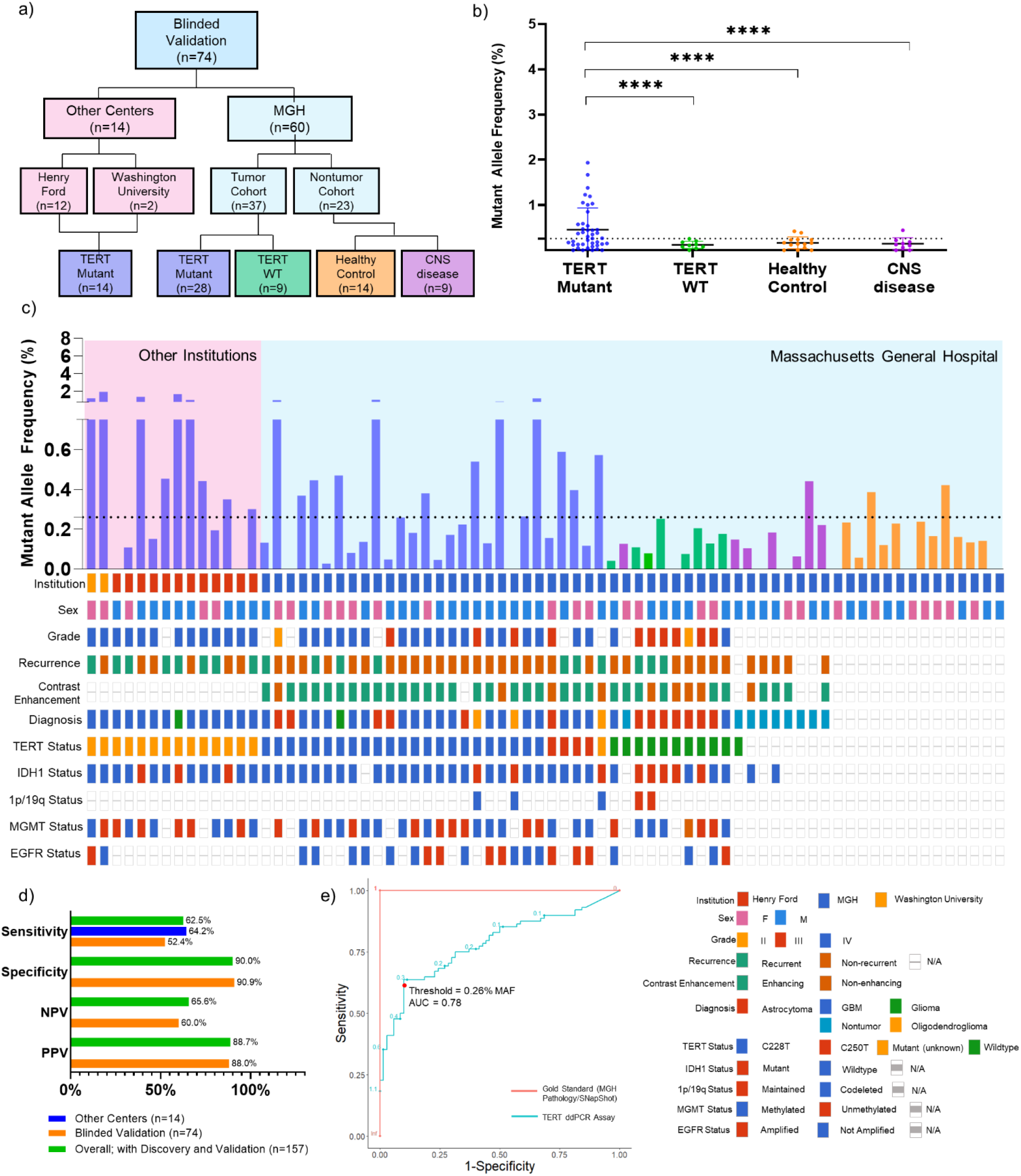
Detection of TERT Promoter Mutation in Plasma of Blinded Multi-Institution Validation Cohort. **(a)** CONSORT diagram depicting patient cohort and design of multi-institution validation cohort. **(b)** cfDNA from 2 mL of matched patient plasma from blinded multi-institution cohort (n=74; n=14 outside institution, TERT Mutant, n=28 MGH TERT Mutant, n=9 MGH TERT WT, n=14 MGH healthy control, n=9 MRI positive non-tumor) was used as input for absolute quantification of TERT mutant and analyzed using the parameters established in the discovery cohort. Data is shown in MAF calculated using the formula described in Methods and Extended Data Fig. 2. MAF for all samples is plotted against sub-classification of patient samples. **(c)** MAF for all TERT Mutant plasma samples are graphed according to sample number, with an accompanying Oncoprint that depicts sample source and genomic landscape. Dotted line indicates threshold of 0.26% MAF used to designate samples as TERT Mutant positive or negative. **(d)** Analytical parameters were calculated from contingency tables and reported. **(e)** ROC Curve depicting change in sensitivity and specificity according to varying threshold. Red point indicates threshold used for analysis, 0.26% MAF. Gold Standard (MGH Pathology/SNapShot) is plotted in red, and TERT ddPCR Assay is plotted in blue.

The blinded multi-institution validation cohort verifies and validates our assay performance for the detection of the TERT promoter mutation in plasma^23-25^. In summary, combining all three cohorts (n=157), we demonstrate a sensitivity of 62.50% (CI, 51.53%-72.60%), a specificity of 90% (CI, 80.48%-95.8%), a PPV of 88.71% (CI, 72.95%-94.17%) and a NPV of 65.62% (CI, 59.04%-71.66) (**Fig. 2e, 3d**). Of patients who were classified positive by plasma analysis but negative by tissue analysis at the MAF threshold selected, the clinical scenario of these false positive samples included 4/34 healthy controls, 1/9 patients with non-glioma CNS disease, and 2/34 TERT WT gliomas.

Within the limits of our cohort, we detected no significant correlation between TERT MAF in plasma and age, duration of symptoms, tumor grade, mutational status (C228T/C250T), tumor volume, contrast enhancement, overall survival and progression free survival (**Extended Data Fig. 4**). However, we noted that patients with contrast enhancing tumors (increased breakdown of blood brain barrier) tended to have higher MAF than patients with non-contrast enhancing tumors (**Extended Data Fig. 1g**). Furthermore, patients with MAF above threshold tended to have poorer progression free survival and overall survival compared to patients with below threshold MAF (**Extended Data Fig. 1a-b**).

We also report perfect concordance in 4 available matched CSF samples from both cohorts compared to a tissue gold standard (TERT mutant n=3; TERT WT n=1). We also detect a TERT mutation in the CSF of a patient sample whose plasma TERT MAF was below the defined assay threshold (**Extended Data Fig. 5**).

### Detection of TERT Promoter Mutations in cfDNA for Longitudinal Monitoring

To assess the performance of the TERT assay in a longitudinal setting, we analyzed cfDNA from serial plasma samples of five patients with TERT mutant glioma maintaining the same analytical parameters. TERT mutant copies over the course of therapy paralleled imaging findings and clinical performance of patients (Figure 4). Each of the patients (excluding P4) had TERT mutant MAF above threshold prior to initial surgical resection and levels returned to baseline postoperatively. On follow up, 3 patients (P1, P2, P3) developed contrast enhancing lesions on MR images after chemoradiation. P2 and P3 had histopathologic confirmation of progression while P1 had clinically diagnosed progression. TERT MAF increased with the development of contrast enhancing recurrent lesions coincident with clinical deterioration. Plasma from P3 was not available for analysis at the time of suspected disease progression. Patients P4 and P5 had stable disease with the TERT mutant levels remaining below baseline over time.

**Figure 4.**
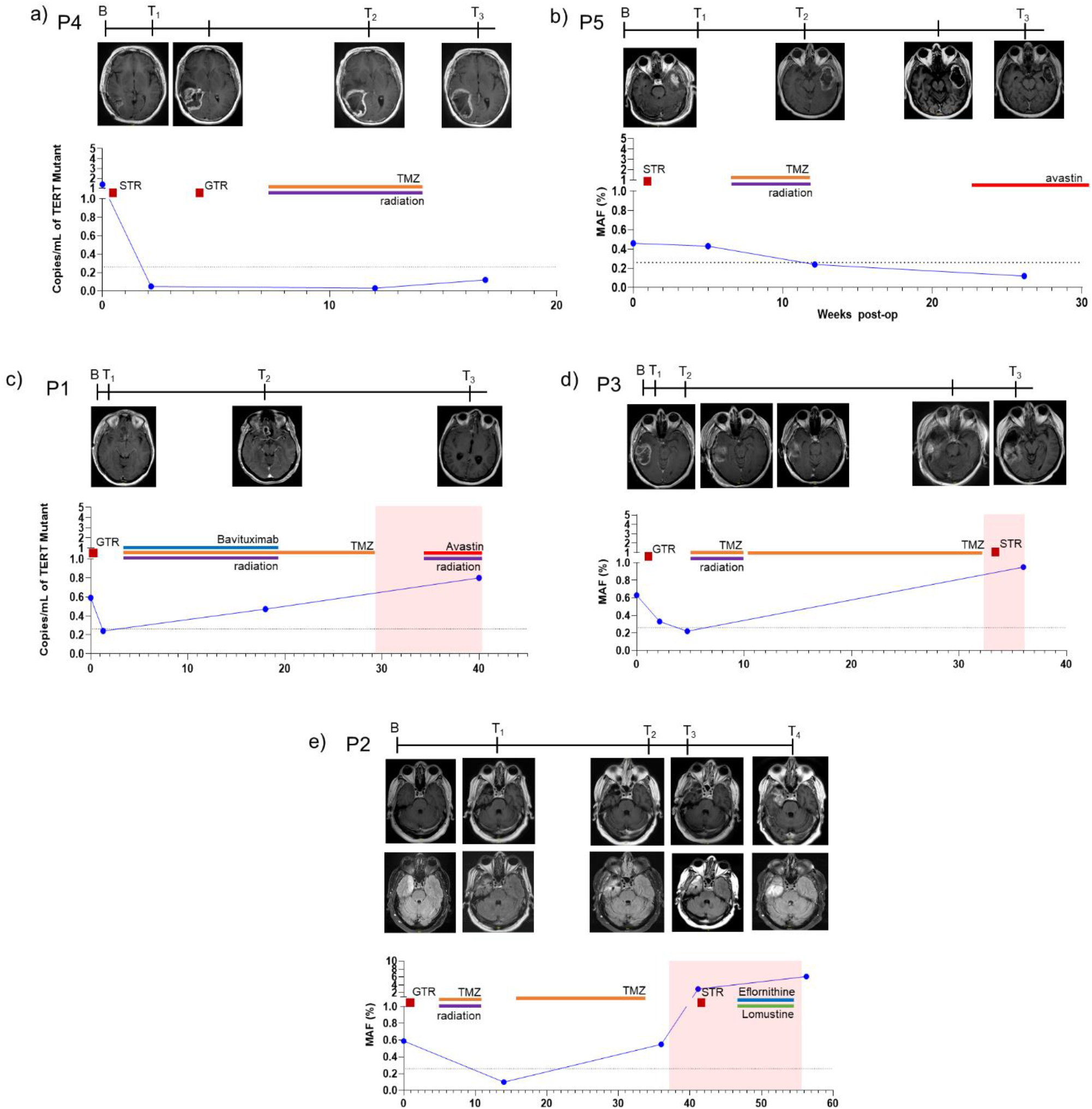
Longitudinal monitoring of TERT promoter mutation in patients with glioma. TERT promoter mutation (copies/mL and MAF) in serial plasma samples obtained from five glioma patients are plotted against time (weeks-post-OP). Cases with stable disease include **(a)** P4 (MGH-19038; grade IV IDH1 wildtype GBM), **(b)** P5 (MGH-19006; grade IV IDH1 wildtype GBM), and progression include **(c)** P1 (MGH-18040; grade IV IDH1 wildtype GBM), **(d)** P3 (MGH-17045, grade IV IDH1 mutant GBM) and **(e)** P2 (MGH-18061; grade III, anaplastic astrocytoma). T1-Weighted, Contrast Enhanced MRI images are provided for timepoints when available. For P2, Axial Flair images are also provided. Timepoints are indicated as baseline (B), timepoint 1 (T1), timepoint 2 (T2), timepoint 3 (T3), and timepoint (T4). Surgical procedures are indicated using a red square (STR=subtotal resection; GTR=gross total resection), orange line indicates administration of chemoradiation and progression is indicated using a red background.

## Discussion

This study demonstrates for the first time, in a majority of glioma patients with a known tissue TERT promoter mutation, the detection of the same mutation in glioma patients’ plasma, using a novel enhanced ddPCR assay. We report an overall sensitivity of 62.5% and a specificity of 90% of detecting cfDNA TERT promoter mutations in plasma compared to a matched tumor tissue gold standard in 114 patients with glioma. The ability to detect the highly prevalent TERT promoter mutation in the plasma of patients with glioma enhances our ability to diagnose, monitor and assess responses to therapy.

In this report, we overcome several challenges that have prevented detection of cfDNA point mutations in specific genes in the plasma of brain tumor patients, including for the TERT promoter specifically. In contrast to other tumor types, where cfDNA point mutations are abundant, this is not the case in glioma. Several prior reports have detected specific oncogenic point mutations in cfDNA, including at the TERT promoter locus, in less than 5% of patients^5,8,26^. This may be related to generally lower levels of CNS-derived circulating tumor DNA (ctDNA) in the blood, thought to be a function of the blood brain barrier^5^. While in our patient cohort, we show that tumor size does not correlate with plasma TERT MAF, (similar to a prior report of non-specific cfDNA quantitation in glioma^27^), we did note a tendency for patients with contrast enhancing lesions (indicative of BBB breakdown) to have higher plasma TERT MAF. Taken with the fact that recurrence timepoints (after surgery and chemoradiation) have more edema and higher plasma TERT MAF compared to baseline despite smaller tumor volumes, we hypothesize that plasma TERT MAF is correlated with breakdown of the blood brain barrier.

Our approach included several technical optimization strategies to detect low concentrations of TERT promoter mutations in plasma. Both C228T and C250T generate identical sequences, which can be detected with a single novel probe with LNA enhancements that stabilize probe-template duplexes to improve SNV discrimination. Furthermore, the TERT promoter region has a high GC-content (>80%) which we address by using 7-deaza-dGTP as a ddPCR additive, which interrupts the formation of these secondary structures. In combination, with standardized handling strategies and an unbiased analytic method we observed a significant improvement in assay sensitivity over previously published reports for TERT promoter detection in plasma of patients with glioma^28^.

This difficulty in detecting cfDNA from brain tumor patients is particularly acute in this population as this challenge also represents a missed opportunity for the liquid biopsy approach in a tumor type where initial or repeat direct tissue sampling may pose substantial neurologic risk. While tissue biopsy will likely remain the gold standard for molecular diagnosis of glioma, the ability to detect oncogenic point mutations in the blood may have specific near-term application in two scenarios: 1) the described TERT promoter mutation assay may be useful for upfront diagnosis. Because some malignant tumors are located in deep or inaccessible locations and would be poor surgical candidates for direct tissue biopsy or resection, the combination of a characteristic set of MRI imaging findings in addition to a liquid biopsy that shows TERT promoter mutation may be sufficient to establish a high positive predictive value for the clinical diagnosis malignant glioma and allow adjunctive treatment with radiation and chemotherapy to proceed with confidence. 2) In another scenario, because of the surgical risk of repeated brain biopsies, a liquid biopsy approach to TERT promoter mutation detection in the blood may be particularly suitable for therapeutic monitoring of disease burden, as exemplified conceptually in the longitudinal cases shown in this report. In our preliminary analysis, levels of TERT promoter mutants, as detected by this assay, do correlate with disease recurrence or progression. This can aid in differentiation between radiation necrosis, pseudoprogression, and true progression, thus minimizing the need for further invasive workup and improving overall quality of care. Furthermore, with emerging insights into intratumoral heterogeneity, the validity of a single, localized tissue biopsy as a true gold standard has begun to be debated^25^, raising the possibility that a liquid biopsy, which effectively samples multiple tumor regions as blood perfuses the solid tumor mass, may offer greater sensitivity for TERT promoter mutation than a single, focal, tissue biopsy, in some patients.

Future studies offer the opportunity for further extension of this particular assay and the general approach of liquid biopsy for brain tumors. The false positive rate of our assay at the threshold chosen was 10%, which is similar to the false positive rate of multimodality imaging analysis to infer diagnosis of gliomas. However, this also included assessment of patients with a very low predicted risk of glioma (e.g. healthy controls with no known MRI abnormalities and patients with fungal abscess/normal pressure hydrocephalus). In practical use, the TERT promoter mutation assay would be preferentially used in situations where disease prevalence is estimated to be high, e.g. in patients with an abnormal MRI scans consistent with glioma, in order to confirm the presence of glioma with a high positive predictive value. Within our cohort, 0/7 patients with an abnormal MRI and non-glioma CNS disease had TERT MAF above threshold. In addition, every patient with abnormal MRI and a plasma TERT MAF above the threshold had a confirmed glioma diagnosis (n=55 tissue TERT Mutant; n=2 tissue TERT WT). This is especially true for patients with contrast enhancing lesions on MRI and plasma TERT MAF above the threshold, as every patient in this subset also had a confirmed glioma diagnosis (n=42 tissue TERT Mutant; n=2 tissue TERT WT). Thus while our assay’s sensitivity compared to a tissue gold standard is encouraging, future studies should assess the positive predictive value of the assay for the overall diagnosis of glial malignancy in the context of abnormal MRI findings, and further, whether the combination of this assay with specific MRI features may also allow for the plasma based diagnosis of specific subtypes of glioma which harbor TERT promoter mutations (e.g. GBM versus anaplastic oligodendroglioma).

While our MAF threshold (0.26%) is optimized based on ROC analysis, from a clinical perspective, it is important to minimize the false positive rate of our assay (10%) below that of multimodal imaging. We plan to reduce this false positive rate by continuing to improve and standardize analytical parameters, and also by exploring the possibility of using different thresholds, and for different clinical and radiographic scenarios. To this end, clinical studies across larger patient cohorts will be required to validate and standardize the gating parameters, plasma volumes and clinical performance of the assay in heterogeneous populations with more extensive longitudinal sampling. Although we used samples from multiple institutions, the samples were heavily weighted (92%) from a single institution, potentially introducing bias in pre-analysis sample handling with concomitant effects on assay performance.

The demonstration herein that the plasma detection of TERT promoter mutations in a majority of glioma patients who harbor these tumor mutations opens the door to extending this to a multi-gene approach to further classify gliomas using a liquid biopsy approach, mimicking tissue based molecular classification. This could be accomplished by careful optimization of assays for common glioma point mutations in the blood such as IDH1 R132H mutations, EGFRVIII mutations, HK23M mutations and others. We also anticipate that this assay will also offer a sensitive method for detecting mutations in plasma cfDNA in other TERT positive tumors.

In conclusion, we demonstrate a novel and highly sensitive ddPCR based TERT promoter mutation assay, utilizing high affinity LNA enhanced probes and the additive 7dG to reduce the formation of secondary structures, for detection and monitoring of TERT promoter mutations (C228T, C250T) in tumor tissue and cfDNA of matched plasma of glioma patients with an overall sensitivity of 62.5% and a specificity of 90% in combined discovery and blinded validation cohorts of 157 samples. The ability to detect TERT mutations, which are highly prevalent in glioma patients, in the plasma enhances our ability to diagnose, monitor and assess response to therapy. Liquid biopsy-based monitoring can significantly impact clinical care by guiding patient stratification for clinical trials, offering new opportunities for the development of targeted therapies ultimately improving patient care.

## Data Availability

Data is available upon request

## Acknowledgements

We would like to thank all of the MGH Neurosurgery clinicians and staff that assisted with the collection of samples. Additionally, we would also like to thank the patients and their families for their participation in this study. We thank Alexandra Beecroft and Abigail Taylor for editorial help. We also thank Alona Muzikansky for help with the statistical analysis and Gur Rotkop and Sharmistha Chakrabortty for help with computational analysis.

## Data/Code Availability Statement

Source data for figures are provided within extended data figures. Code is available on GitHub Repository koushikmuralidharan/BI LL.

## Author Contributions

L.B., B.S.C. conceptualized and supervised the study and designed the experiments; K.M., extracted cfDNA and performed ddPCR experiments. A.Y., extracted cfDNA from plasma samples. J.S.,and Z.R., consented patients, collected specimens and the relevant clinical data. K.K., and L.W., extracted tumor DNA. S.L., optimized the primer sequence. H.Z., H.L assisted with biostatistics and use of mathematical modeling for data analysis. C.B., M.R.C., S.N.K, G.M.S, B.V.N, W.T.C, P.S.J., and D.P contributed to study design, collection of samples, and manuscript preparation. K.M., L.B., and B.S.C., analyzed all data, generated figures and wrote the paper with input from all authors.

## Methods

### Study population

The study population (n=157) included patients 18 years or older with pathology confirmed TERT mutant or wildtype gliomas who underwent surgery at either the Massachusetts General Hospital (MGH), Henry Ford Healthy System (HFHS), or at Washington University St. Louis (WUStL) and age-matched healthy controls (**Figure 2a; Table 1**).The study population was divided into a discovery cohort (n=83; 46 TERT Mutant, 17 TERT wild-type, 20 matched healthy controls) and a blinded multi-institution validation cohort (n=74; 42 TERT Mutant, 9 TERT WT, 14 matched healthy controls, 9 non-tumor MRI positive). For glioma cohorts, exclusion criteria consisted of history of other primary or metastatic cancers, active infectious disease, current or previous enrollment in clinical trials, and hemolyzed plasma samples. All healthy control subjects were screened for pertinent oncologic and neurologic medical histories. Individuals with a history of cancer, neurological disorders, and infectious diseases were excluded from the study. All samples were collected with informed consent under Partners institutional review board (IRB)-approved protocol number 2017P001581. BRISQ guideline reports are included in **Supplementary Table 1**. Samples are taken from patient population undergoing treatment at MGH, HFHS, or WUStL. Some samples may still be available and could be shared with appropriate documentation (IRB, MTA etc.). ExoLution PLUS is a proprietary kit available from Exosome Diagnostics (a BioTechne brand).

### Tumor tissue processing

Tumor tissue was microdissected and suspended in RNAlater (Ambion) or flash-frozen and stored at −80°C.

### Patient plasma processing

Whole blood was collected using K2 EDTA tubes with an inert gel barrier (BD Vacutainer Blood Collection Tubes). Within 2 hours of collection, samples were centrifuged at 1,100 xg for 10 minutes at 20°C to separate the plasma from the hematocrit and filtered using 0.8 μm filters. 1 ml aliquots were stored at −80°C for later downstream analysis. With the exception of the longitudinal samples, all baseline samples were collected prior to surgical resection.

### Cell lines

Human carcinoma cell line A431 (ATCC CRL-1555) was cultured in Dulbecco’s modified essential medium with high glucose (DMEM; Gibco, Invitrogen Cell Culture), containing 10% fetal bovine serum (FBS; Life Technologies Corporation) and 1% Penicillin/Streptomycin (Life Technologies). Human glioma cell line U87 (ATCC HTB-14) was cultured in DMEM with high glucose, containing 10% FBS and 1% Penicillin/Streptomycin. Human brain microvascular endothelial cells (HBMVEC) were kindly provided by Xandra O. Breakefield and cultured using endothelial basal medium (EGM-2 MV Microvascular Endothelial Cell Growth Medium-2 BulletKit, Lonza). All cell lines were grown to 50-70% confluency prior to gDNA extraction to minimize cell death and optimize quality of gDNA. All cell lines were verified monthly for mycoplasma contamination using commercial mycoplasma PCR (Mycoplasma PCR Detection Kit, Applied Biological Materials) to ensure lack of mycoplasma contamination.

### DNA isolation

DNA was isolated from cell lines and frozen tumor tissue using the DNeasy Blood and Tissue Kit (Qiagen) as recommended by the manufacturer. DNA was eluted in AE buffer (Qiagen) and stored at −20°C until further processing. DNA concentration and purity were determined using the NanoDrop One (ThermoFisher Scientific).

### Plasma cfDNA isolation

Circulating nucleic acid was extracted from plasma using the QIAamp Circulating Nucleic Acid Kit (Qiagen) or ExoLution PLUS (Exosome Diagnostics) as per the manufacturer’s instructions. cfDNA eluted in 20 μL AVE buffer (QIAamp Circulating Nucleic Acid Kit) or in 20 μL nuclease-free water (ExoLution PLUS Kit) and stored at −20°C until quantification and subsequent ddPCR test.

### TERT ddPCR assay

Since both the C228T and C250T TERT promoter mutations yield the same sequence (Figure 1b), a single probe was used to detect both mutations. A second probe was also used to recognize the C228 wild type locus. As described by McEvoy et. al, Locked Nucleic Acid (LNA) modifications were introduced on probes due to the short size of the probe, indicated by “+”^19^. The sequences for the probes used are as follows: TERT promoter mutant (5’-FAM/CCC +C+T+T+CCGG/3IABkFQ/) and TERT promoter wild type (5’-HEX/CCC C+C+T +CCG G/3IABkFQ/). Probes were synthesized by Integrated DNA Technologies (IDT). ddPCR amplification was performed using either 4 μL of cfDNA template or using 100 ng of tumor gDNA, 1x ddPCR Supermix for probes (no dUTP, Bio-Rad), either 1x Q-sol or 200 mM 7-deaza-dGTP (7dG; New England Biolabs), 250 nM of each probe and 900 nM of each primer (5’-CCTGCCCCTTCACCTTCCAG-3’ and 5’-AGAGCGGAAAGGAAGGGGA-3’) with template (100 ng of tumor tissue or 4 uL of cfDNA) in a total reaction mix of 20 μL. The QX200 manual droplet generator (Bio-Rad) was used to generate droplets. Thermocycling conditions were as follows: 95°C (51% ramp) for 10 minutes, 40 cycles of 94°C (51% ramp) for 30 seconds and 57°C for 1 minute, followed by 98°C for 10 minutes and held at 4°C until further processing. Droplets were counted and analyzed using the QX200 droplet reader (Bio-Rad) and QuantaSoft analysis (Bio-Rad) was performed to acquire data.

### Quantification of Copies/mL of Plasma

Copies per mL of plasma is calculated by taking copies/20μL (C; provided by QuantaSoft), multiplied by elution volume in μL (EV), divided by the total volume added to the reaction (TV), divided by the plasma volume (PV). In short, Copies/mL of plasma = C^*^EV/TV/PV.

### R-based ddPCR Analysis of MAF from Plasma Samples

Gates were constructed for both channel 1 (4-8) and channel 2 (2-6) in increments of 0.05. Combinations of channel 1 gates and channel 2 gates were used to calculate MAF, the number of channel 1 positive droplets divided by the number of channel 2 positive droplets. Thresholds were calculated to fulfill one of the following criteria: maximize specificity, set specificity close to 90%, and to minimize the distance from the ROC curve to the point (0,1). These gating strategies (trained using discovery cohort) were then used to analyze the data from the multi-institution validation cohort in a blinded fashion. Code available on GitHub Repository koushikmuralidharan/BILL.

### Statistical Analysis

Statistical analyses were performed using unpaired two-tailed Student’s *t*-test in GraphPad Prism 8 software and *p* < 0.05 was considered statistically significant. Confidence intervals were calculated using exact binomial distributions. The results are presented as the mean ± SD. “***” indicates p-value less than or equal to 0.001, and “****” indicates p-value less than or equal to 0.0001.

## References

1. Louis, D. N. et al. The 2016 World Health Organization Classification of Tumors of the Central Nervous System: a summary. Acta Neuropathol. 131, 803–820 (2016).

2. Wang, Y, et al. Detection of tumor-derived DNA in cerebrospinal fluid of patients with primary tumors of the brain and spinal cord. Proc. Natl. Acad. Sci. U. S. A. 112, 9704–9709 (2015).

3. Chen, W. W. et al. BEAMing and Droplet Digital PCR Analysis of Mutant IDH1 mRNA in Glioma Patient Serum and Cerebrospinal Fluid Extracellular Vesicles. Mol. Ther. Nucleic Acids 2, e109 (2013).

4. Miller, A. M. et al. Tracking tumour evolution in glioma through liquid biopsies of cerebrospinal fluid. Nature 565, 654–658 (2019)

5. Bettegowda, C, et al. Detection of circulating tumor DNA in early- and late-stage human malignancies. Sci Transl. Med. 6, 224ra24 (2014).

6. Shankar, G. M. et al. Rapid Intraoperative Molecular Characterization of Glioma. JAMA Oncol 1, 662–667 (2015).

7. Zill, O. A. et al. The Landscape of Actionable Genomic Alterations in Cell-Free Circulating Tumor DNA from 21,807 Advanced Cancer Patients. Clin. Cancer Res. 24, 3528–3538 (2018).

8. Piccioni, D. E. et al. Analysis of cell-free circulating tumor DNA in 419 patients with glioblastoma and other primary brain tumors. CNS Oncol 8, CNS34 (2019).

9. Eckel-Passow, J. E. et al. Glioma Groups Based on 1p/19q, IDH, and TERT Promoter Mutations in Tumors. N. Engl. J. Med. 372, 2499–2508 (2015).

10. Labussière, M, et al. TERT promoter mutations in gliomas, genetic associations and clinico-pathological correlations. British Journal of Cancer vol. 111 2024-2032 (2014).

11. Colebatch, A. J. et al. Optimizing Amplification of the GC-Rich TERT Promoter Region Using 7-Deaza-dGTP for Droplet Digital PCR Quantification of TERT Promoter Mutations. Clin. Chem. 64, 745–747 (2018).

12. Bell, R. J. A. et al. Understanding TERT Promoter Mutations: A Common Path to Immortality. Mol. Cancer Res. 14, 315–323 (2016)

13. Colebatch, A. J., Dobrovic, A, & Cooper, W. A. TERT gene: its function and dysregulation in cancer. J Clin. Pathol. 72, 281–284 (2019).

14. Killela, P. J. et al. TERT promoter mutations occur frequently in gliomas and a subset of tumors derived from cells with low rates of self-renewal. Proc. Natl. Acad. Sci. U. S. A. 110, 6021–6026 (2013).

15. Vinagre, J, et al. Frequency of TERT promoter mutations in human cancers. Nat. Commun. 4, 2185 (2013).

16. Kang, S,, Ohshima, K,, Shimizu, M,, Amirhaeri, S, & Wells, R. D. Pausing of DNA synthesis in vitro at specific loci in CTG and CGG triplet repeats from human hereditary disease genes. J. Biol. Chem. 270, 27014–27021 (1995).

17. Motz, M,, Pääbo, S, & Kilger, C, Improved Cycle Sequencing of GC-Rich Templates by a Combination of Nucleotide Analogs. BioTechniques vol. 29 268-270 (2000).

18. Mamedov, T. G. et al. A fundamental study of the PCR amplification of GC-rich DNA templates. Comput Biol. Chem. 32, 452–457 (2008).

19. McEvoy, A. C. et al. Sensitive droplet digital PCR method for detection of TERT promoter mutations in cell free DNA from patients with metastatic melanoma. Oncotarget 8, 78890–78900 (2017).

20. Corless, B. C. et al. Development of Novel Mutation-Specific Droplet Digital PCR Assays Detecting TERT Promoter Mutations in Tumor and Plasma Samples. J. Mol. Diagn. 21, 274–285 (2019).

21. Attali, D,, Bidshahri, R,, Haynes, C, & Bryan, J, ddpcr: an R package and web application for analysis of droplet digital PCR data. F1000Research vol. 5 1411 (2016).

22. Zheng, Z, et al. Anchored multiplex PCR for targeted next-generation sequencing. Nat. Med. 20, 1479–1484 (2014).

23. Juratli, T. A. et al. Intratumoral heterogeneity and promoter mutations in progressive/higher-grade meningiomas. Oncotarget 8, 109228–109237 (2017).

24. Nakajima, N, et al. BRAF V600E, TERT promoter mutations and CDKN2A/B homozygous deletions are frequent in epithelioid glioblastomas: a histological and molecular analysis focusing on intratumoral heterogeneity. Brain Pathol. 28, 663–673 (2018).

25. Sottoriva, A, et al. Intratumor heterogeneity in human glioblastoma reflects cancer evolutionary dynamics. Proc. Natl. Acad. Sci. U. S. A. 110, 4009–4014 (2013).

26. Juratli, T. A. et al. TERT Promoter Mutation Detection in Cell-Free Tumor-Derived DNA in Patients with IDH Wild-Type Glioblastomas: A Pilot Prospective Study. Clinical Cancer Research vol. 24 5282-5291 (2018).

27. Nørøxe, D. S. et al. Cell-free DNA in newly diagnosed patients with glioblastoma - a clinical prospective feasibility study. Oncotarget 10, 4397–4406 (2019).

28. Juratli, T. A. et al. Promoter Mutation Detection in Cell-Free Tumor-Derived DNA in Patients with Wild-Type Glioblastomas: A Pilot Prospective Study. Clin. Cancer Res. 24, 5282–5291 (2018).

29. Gao, J, et al. Integrative analysis of complex cancer genomics and clinical profiles using the cBioPortal. Sci. Signal. 6, l1 (2013).

30. Cerami, E, et al. The cBio cancer genomics portal: an open platform for exploring multidimensional cancer genomics data. Cancer Discov. 2, 401–404 (2012).

